# LEVELS OF EXCLUSIVE BREASTFEEDING AND CORRELATES AT 10WEEKS POST-DELIVERY IN A RURAL SETTING

**DOI:** 10.1101/2024.04.15.24305842

**Authors:** Adenike Oluwakemi Ogah, James-Aaron Ogah, Elizabeth Edigwu Ogah

## Abstract

**Background:** Sustaining a high rate of adherence to exclusive breastfeeding may be challenging in resource constrained setting. This study established the levels of exclusive breastfeeding, alternate feeding patterns, health and growth impacts, among 10 weeks old infants in a remote, understudied village in East Africa.

**Subject and methods:** This was a secondary cross-sectional analysis of a prospective cohort data. Data of 457 out of the 529 mother-newborn pairs recruited from birth were obtained and analysed at 10 weeks post-delivery in the postnatal clinic. Infant feeding patterns were recorded and anthropometry were measured and assessed using NICE criteria for static weight growth. Mothers were interviewed using the Edinburgh postpartum depression score to assess emotional status. Both parametric and non-parametric statistics and binary logistic regression model were applied to examine the relationship between maternal/infant characteristics, feeding patterns and infant weight growth. The results were presented in p-values, Odds ratio and 95% confidence interval.

**Results:** Clinic attendance had declined below and prevalence of infant weight faltering had increased above the 6 weeks records. The prevalence of infant weight faltering was 3.7% on NICE criteria, higher than the 2.8% with WHO criteria, but there was 93% concordance between the 2 growth assessment tools. Exclusive breastfeeding rate was still high at 96.3%, however, 10.3% of these infants were being breastfed infrequently. The 3 infants that were not exclusively breastfed were offered infant formula (1) using bottle, maize (1) and millet porridge (1) using cup. Infrequent breastfeeding was the major predictor of infant weight faltering, OR 5.59; 95%CI 1.94, 16.14; p=0.001. For every unit increase in frequency of breastfeeding, the infant weight-for-age z-score is likely to increase by 0.15 z-score. To maintain the infant weight z-score at 0 (median), baby should be breastfed on the average of 10times a day consistently. The head circumference and then length were more deficient than the weight in this cohort, despite high level of EBF rate. Maternal depression score was high in the groups of infants that were adequately breastfed and weight thriving, compared to their counterparts.

**Conclusion:** Both the practice of EBF and the daily frequency of breastfeeding should be closely monitored; infant head circumference and length assessments should be integrated into the routine growth monitoring programs in these rural MCH clinics. Lactational and mental support programs should be strengthened in the rural MCH systems, to assist mothers to achieve full exclusive breastfeeding easily. The factors contributing to decline in clinic visits need to be identified and mitigated. Home visits should be intensified for infants lost to follow up.

## Background

Exclusive breastfeeding (EBF) is the best feeding practice for infants aged from birth to 6 months because breastmilk supplies all the vital nutrients required for optimal growth and development during this period. EBF has faced challenges of adherence, despite information of its importance being given to mothers. This poor adherence exposes the infant to a barrage of health problems such as diarrhea^1^

Even though, more than 95% of infants in Africa, are currently breastfed, feeding practices are often inadequate and include offering water, and other liquids, to breastfed infants. Consequently, the rate of exclusive breast-feeding is low despite promotion efforts deployed by WHO and Unicef, local governments, and non-governmental organizations. The rate of bottle-feeding is high in some countries (exceeding 30% in Tunisia, Nigeria, Namibia and Sudan)^2^

It has come to global public health attention, that a significant number of breastfeeding mothers of infants <6 months might be facing challenges in breastfeeding.^3^ In a systematic review of barriers to exclusive breastfeeding in low and middle income countries, noted that up to one-third of mothers in Pakistan, Nigeria and Ghana discontinued exclusive breast-feeding because they felt it was a stressful, frustrating and/or painful experience, as a result of illness or breast problems.^4^ Urbanization and mothers’ education also tend to shorten breastfeeding. For instance, a Ghana study, among city-dwelling professional mothers, documented EBF rate at six months to be as low as 10.3 %.^5^

Incomplete exclusive breastfeeding has both short and long term adverse consequences on infant, mother, family, community, nation and globally. Hence, this study was designed to determine the levels of EBF, risk factors, alternative feeding patterns and weight growth profiles among 457, 10 weeks old infants attending MCH clinic in a rural East African community.

### Concept of the study

Figure 1, shows the maternal and neonatal determinants of infant weight faltering that were investigated in this study.

**Fig. 1:**
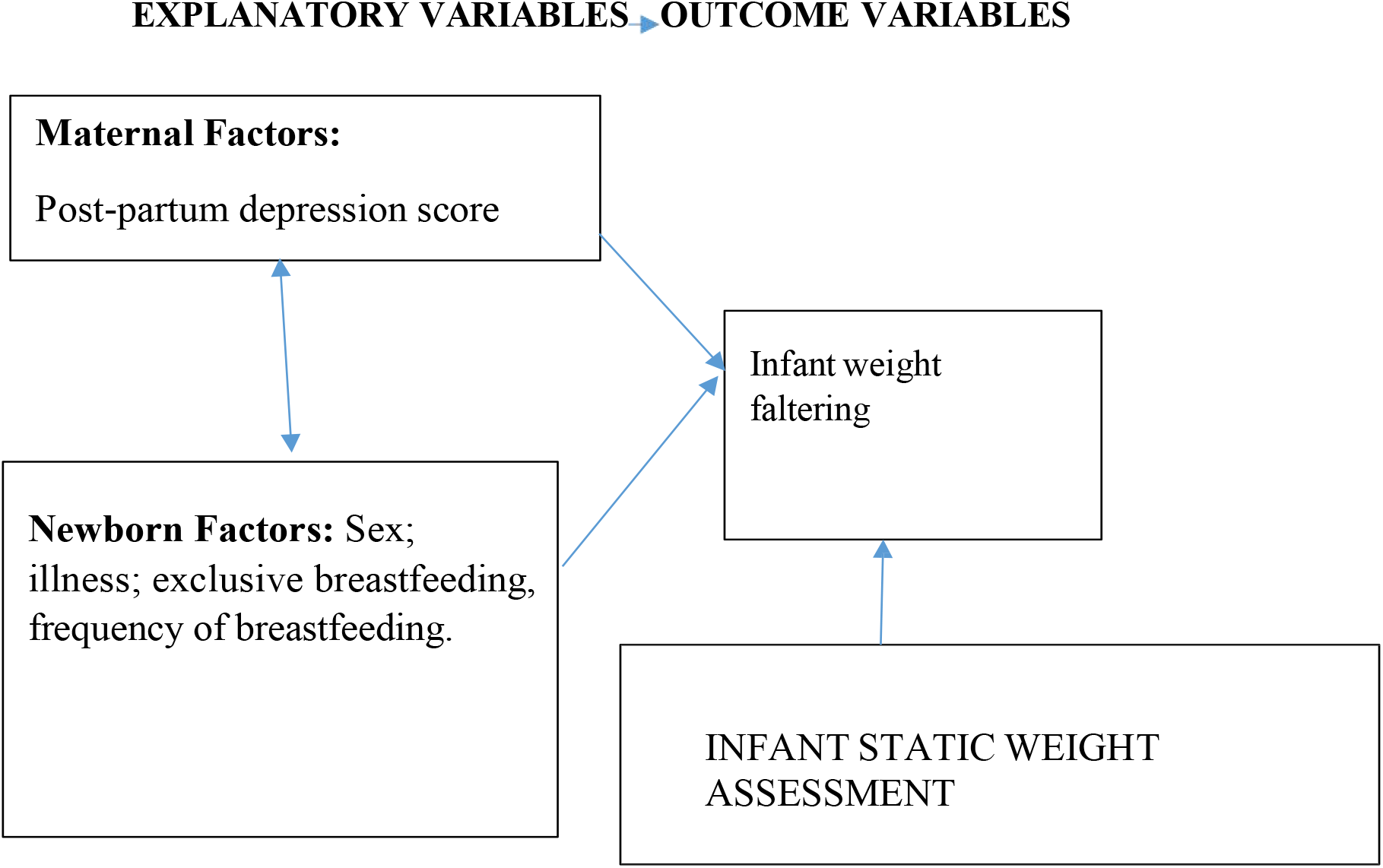
Concept of the study.

## Materials and methods

The methods employed in carrying out this study is discussed in this section.

### Study setting

Rwanda is a low-income, agricultural and landlocked country with approximately 11 million people living in five provinces, covering an area of 26,338 km.^6^ It is called the home of a ‘1000 hills’^6^. The limited area of flat land available in most part of Rwanda is a hindrance to farming, animal rearing and construction of standard residential houses, among others. For example, the recommended minimum of 50 feet distance between source of household water and sewage tank in residential yards are often compromised during construction, leading to contamination of water source. There are 2 peak raining seasons in the country: April to June and September to November.

Rwanda has an average of 4.4 persons per household and a gross domestic product per capita of US $780.80. About half (48%) of its population is under 19 years of age and 39% live below the poverty line with a life expectancy at birth of 71.1 years for women and an adult literacy rate of 80% among 15–49 years old women. In addition, 87.3% of the population have health insurance and access to health services; spending an average of 47.4 min to reach a health centre.

Non-availability of regular supplies of clean and safe water, has been a longstanding problem in Rwanda, probably as a result of its landlocked and hilly terrains, making construction and supply of piped water, a major challenge. The public piped water flow infrequently and the taps and pipes may be rusted and breached in some places, especially in the rural areas, further contributing to contamination of household water. Many families store rain water in big tanks for use in their homes and this may become polluted (in the writer’s opinion) because of difficulties of cleaning these storage tanks. A few non-profit organizations, such as USAID and Water-for-life have sunk bore holes in strategic locations in a few number of villages in the country, with the aim of alleviating this water problem.

Gitwe village is located on a high altitude of 1,674 meters above sea level, in the southern province, 240km from Kigali, which is the capital city of Rwanda. Gitwe general hospital began in 1995, immediately after the genocide, for the purposes of providing medical services and later training to this isolated community. The hospital currently has 100% government support, since year 2020. The maximum number of deliveries at the hospital per month was about 200 and about 50 infants visit the maternal and child health clinic (MCH) per day. There are only 2 immunisation days per week.

Some of the challenges in the hospital include poor specialist coverage and few trained health workers, poor supply of equipment, water, electricity, laboratory services and medicines. Challenging cases are referred to the University of Rwanda Teaching Hospital in Butare or Kigali. Gitwe village was selected for this study, because there was no published birth data from this poorly researched, remote, relatively unaccessible community. In 2019, birth and up to 9 months of age feeding and growth data on 529 healthy mother-singleton newborn pairs were compiled in this village, over a period of 12 months in the delivery, postnatal wards and clinics of Gitwe general hospital and at its annex, the MCH.^6^

### Data source and sample

This was a secondary analysis of data collected for a prospective cohort study on growth faltering over a period of 12 months, until infant was 9 months old. Sample size of 529 was calculated from the Epitool sample size calculator.^7^ Mother-newborn pairs were recruited consecutively, on first-come-first-serve basis at birth. Maternal file review and newborn anthropometry [weight (kg), length (cm) and head circumference (cm) measurements, their z-scores and percentiles, were recorded to the nearest decimals] were carried out, soon after birth. Newborn gestational age was determined using the record of the maternal first date of the last menstrual period (LMP), fetal ultrasound dating and/or expanded new Ballard criteria. Weight faltering at 10 weeks post-delivery was defined by NICE static weight assessment of weight-for-age z-score of ≤2.^8^ Weight-for-age z-score at 10 weeks was obtained from PediTool calculator, which was based on WHO growth standards and used the infants’ corrected age.^9^ Mothers were interviewed using standardized infant feeding questionnaires^10^ and the Edinburgh postpartum depression questionnaire/score^11^ at 10 weeks post-partum in the postnatal clinic. The questionnaires were read to the mothers and filled by the research assistants.

To ensure the quality of data collected, 2 registered nurses were trained as research assistants at Gitwe Hospital on the over-all procedure of mother and newborn anthropometry and data collection by the investigator. The questionnaires were pre-tested before the main data collection phase, on 10 mother-infant pair participants (2% of the total sample). The investigator closely followed the day-to-day data collection process, to ensure completeness and consistency of the questionnaires administered each day, before data entry.

### Statistical analysis

Data clean up, cross-checking and coding were done before analysis. These data were entered into Microsoft Excel statistical software for storage and then exported to Statisty-free web-based online statistics app^12^ for further analysis. Both descriptive and analytical statistical procedures were utilized. Participants’ categorical characteristics were summarised in frequencies and percentages. Chi test was used to determine association between mother/infant characteristics and weight faltering. Numerical characteristics such as maternal depression scores were presented in median and interquartile ranges for skewed data. Non-parametric (for skewed data) and parametric (for normally distributed data) tests were performed to determine relationship between variables and infant weight faltering. Binary logistic regression models were created to identify the predictors of infant weight faltering and to generate the odds ratio. Factors with p-values <0.1 were included in the regression model. Odds ratio (OR), with a 95% confidence interval (CI) were computed to assess the strength of association between independent and dependent variables. For all, statistical significance was declared at p-value <0.05. The reporting in this study were guided by the STROBE guidelines for observational studies.^13^

### Ethics

The Health Sciences Research Ethics Committee of the University of the Free State in South Africa gave the ethical approval for the collection of the primary data for the original study-‘Growth and growth faltering in a birth cohort in rural Rwanda: a longitudinal study’ with Ethical Clearance Number: UFS-HSD2018/1493/2901. Written permission to collect data was obtained from the Director of Gitwe Hospital, Rwanda. Verbal consent were obtained from eligible mothers as they were recruited consecutively from the postnatal ward after delivery, with the attending nurse as a witness. The content of the research information booklet and consent letter were read out to the mothers and their permission was sought after ensuring that they understood the purposes, methods, pros and cons of the research. The participants were given research identity numbers and the principal investigator was responsible for the safe keeping of the completed questionnaires and collected data, to ensure anonymity and confidentiality of the participants.

## Results

The following are the results obtained from the study.

### Participants

Five hundred and ninety-seven (597) babies were delivered at Gitwe Hospital, Rwanda, between 22^nd^ January and 9^th^ May 2019, out of which, eligible 529 mother-newborn pairs were enrolled into the prospective cohort study at birth, Figure 2. 512 of these 529 babies were assessed at 6 weeks postnatal age at the Maternal & Child Health Clinic between 5th March 2019 and 20th June 2019. Between 2^nd^ of April and 18^th^ July 2019, 467 babies from this cohort were reviewed again at 10weeks post-delivery.

**Fig 2:**
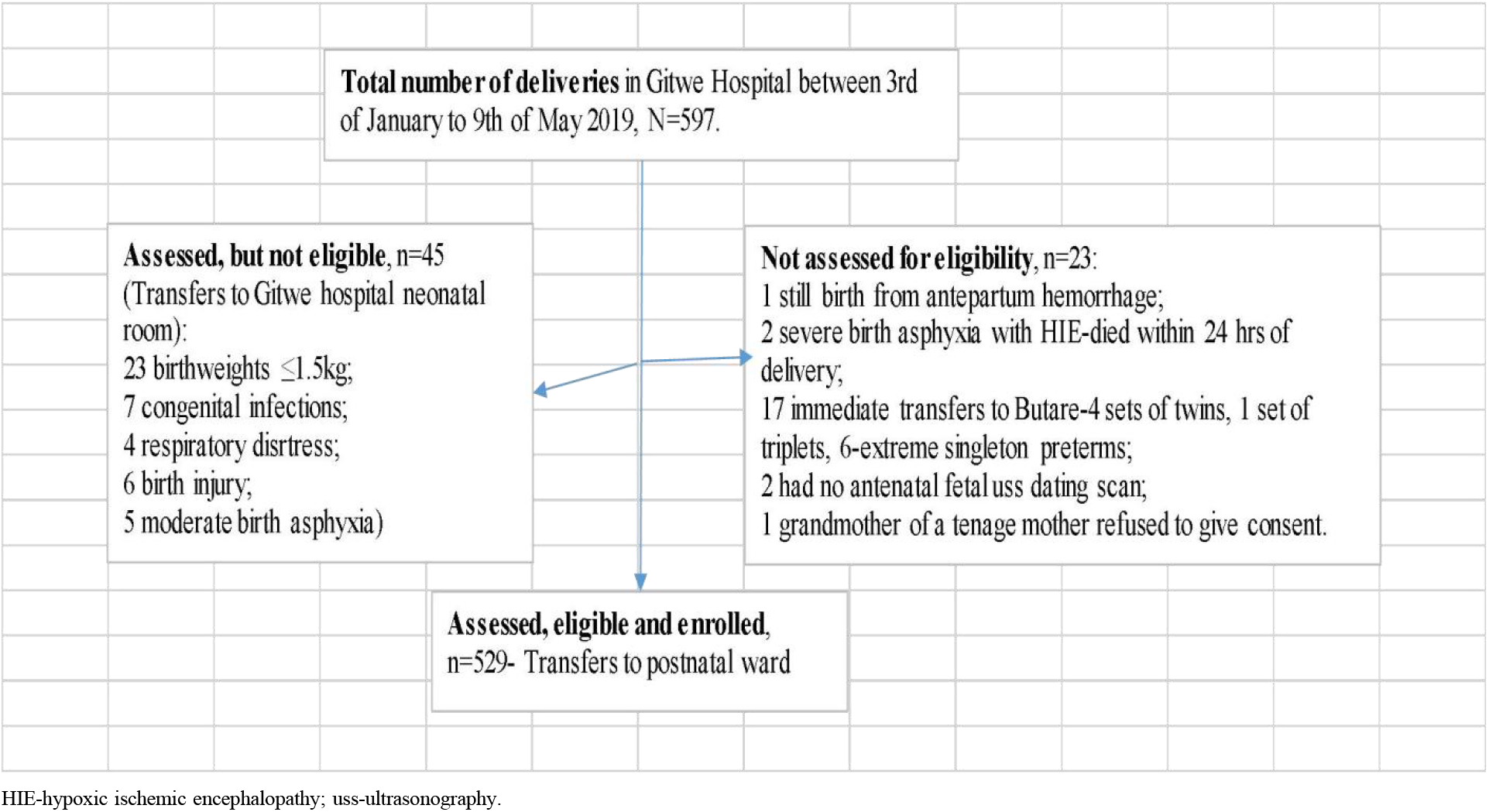
Flow of participants from admission to recruitment into study.

Growth and feeding data were collected from a total of 467 babies at 10 weeks post-delivery. 62 out of the 529 (11.7%) babies recruited from birth were absent at the 10weeks clinic. Ten babies were further excluded from analysis due to incomplete data. Hence, data from a total of 457 babies were analysed: 214 girls (46.8%) and 243 boys (53.2%) at the 10 weeks postnatal clinic. Their median age was 10weeks; mean age was 8.47weeks (sd 3.52). All the 457 (100%) babies were still being breastfed and 96.3% were exclusively breastfed. Two boys and 1 girl were no longer exclusively breastfed. One of the boys, that were no longer on exclusive breastfeeding, received infant formula milk with bottle. The remaining 2 were fed with millet and maize porridge using cup and spoon. Median maternal depression score was 7 (IQR 3,8).

### Weight faltering according to NICE and WHO criteria at 10 weeks post-delivery

Overall, 17 (3.72%) out of the 457 babies were weight faltering according to NICE static assessment. According to WHO criteria, 13(2.84%) were weight faltering, 396 (86.7%) were normal and 48 (10.5%) were overweight. Only 4 (23.5%) infants out of the 17 weight faltering showed clinical signs of wasting.

A Fisher exact test was performed between NICE.weight assessment and WHO.weight assessment. There was a statistically significant relationship between NICE.weight assessment and WHO.weight assessment, p = <.001. The Pearson correlation between NICE.weight assessment and WHO.weight assessment was very strong at 0.93

### Infant characteristics and weight faltering

A higher percentage of boys (5.3%) compared to girls (1.9%); and poorly fed babies (12.8%) compared to adequately fed (2.7%) were weight faltering in Table 2. Of note, there was no significant difference(p=0.206) in the frequency of breastfeeding between the baby girl (mean frequency in 24hrs was 9.7, sd 1.9) and boy (mean frequency in 24hrs was 9.92, sd 1.8).

**Table 1:** Crosstabulation of infant weight assessments based on NICE and WHO criteria, n=457.

|  |  | WHO weight criteria |  |  |
| --- | --- | --- | --- | --- |
|  |  | Weight faltering | Not weight faltering | Total |
| NICE.weight criteria | Weight faltering | 13 | 4 | 17 |
|  | Not weight faltering | 0 | 440 | 440 |
| Total |  | 13 | 444 | 457 |

**Table 2:**
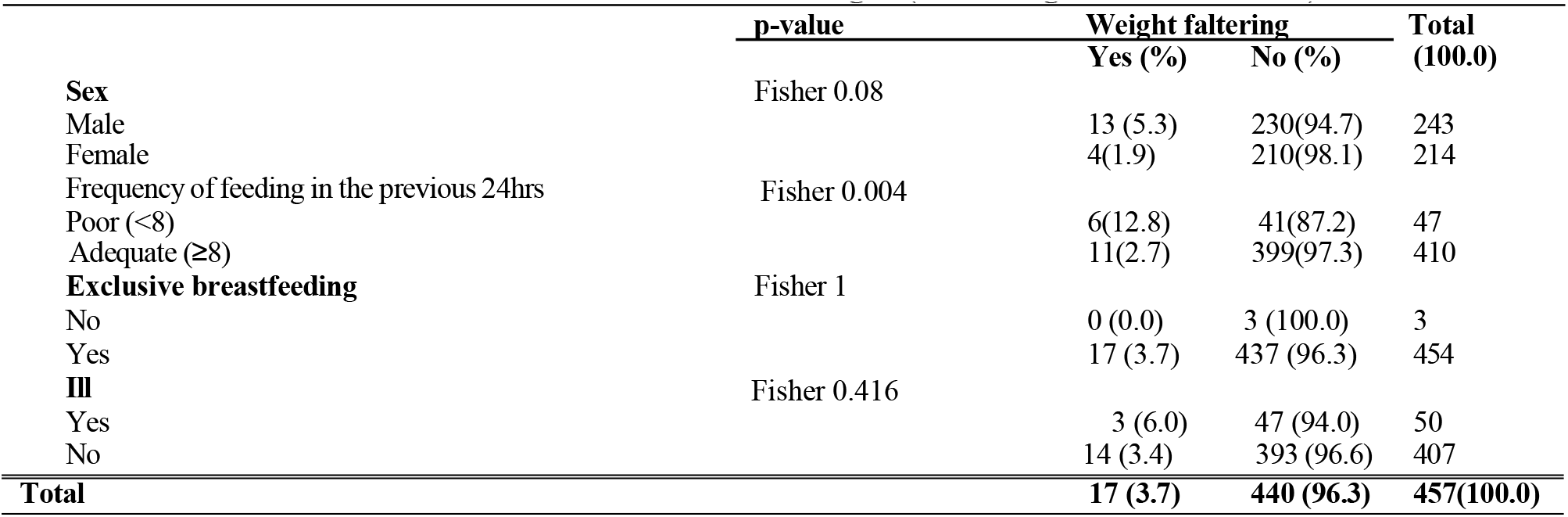
Crosstabulation of infant characteristics and weight (according to NICE criteria), n=457.

|  | p-value | Weight faltering |  | Total (100.0) |
| --- | --- | --- | --- | --- |
|  |  | Yes (%) | No (%) |  |
| **Sex** | Fisher 0.08 |  |  |  |
| Male |  | 13 (5.3) | 230(94.7) | 243 |
| Female |  | 4(1.9) | 210(98.1) | 214 |
| **Frequency of feeding in the previous 24hrs** | Fisher 0.004 |  |  |  |
| Poor (<8) |  | 6(12.8) | 41(87.2) | 47 |
| Adequate ($\geq 8$) | | 11(2.7) | 399(97.3) | 410 |
| **Exclusive breastfeeding** | Fisher 1 |  |  |  |
| No |  | 0 (0.0) | 3 (100.0) | 3 |
| Yes |  | 17 (3.7) | 437 (96.3) | 454 |
| **Ill** | Fisher 0.416 |  |  |  |
| Yes |  | 3 (6.0) | 47 (94.0) | 50 |
| No |  | 14 (3.4) | 393 (96.6) | 407 |
| **Total** |  | **17 (3.7)** | **440 (96.3)** | **457(100.0)** |

### Infant feeding, weight assessment and maternal depression score

Descriptive statistics showed that, the groups 2 infants (non-weight faltering and adequate frequency of breastfeeding) were associated with higher values of maternal depression score (Mdn = 7) than groups 1 infants (weight faltering and inadequate frequency of breastfeeding; Mdn = 0). Mann-Whitney U-Test was statistically significant, [U = 1845, n1 = 440, n2 = 17 p = <.001 for weight. U = 2555.5, n1 = 47, n2 = 410 p <.001 for frequency of feeding. The effect sizes of the weight variable was small; while that of the frequency of feeding was medium, r= 0.17 and 0.4, respectively.

### Logistic regression analysis of male sex and poor feeding predicting weight faltering according to NICE criteria

Logistic regression analysis was performed to examine the influence of Sex-male and Frequency of feeding-poor on variable NICE.Weight to predict the value “weight faltering”. Logistic regression analysis showed that the model as a whole was significant (Chi2(2) = 12.47, p .002, n = 457).

The coefficient of the variable Sex-male was b = 1.14, which was positive. This meant that if the value of the variable is Sex-male, the probability of the dependent variable being “weight faltering” increases. However, the p-value of .052 indicates that this influence is not statistically significant. The odds ratio of 3.13 means that if the variable is Sex-male, the probability that the dependent variable is “weight faltering” increases by 3.13 times.

The coefficient of the variable Frequency of feeding-poor was b = 1.72, which was positive. This meant that if the value of the variable is Frequency of feeding-poor, the probability of the dependent variable being “weight faltering” increases. The p-value of 0.001 indicates that this influence was statistically significant. The odds ratio of 5.59 meant that if the variable is Frequency of feeding-poor, the probability that the dependent variable is “weight faltering” increases by 5.59 times, Table 3.

**Table 3:** Logistic regression model of male sex and poor feeding predicting weight faltering.

|  | Coefficient B | Standard error | z | p | Odds Ratio | 95% conf. interval |
| --- | --- | --- | --- | --- | --- | --- |
| Constant | -4.35 | 0.55 | 7.93 | <.001 | 0.01 | 0 - 0.04 |
| Sex-Male | 1.14 | 0.59 | 1.95 | .052 | 3.13 | 0.99 - 9.91 |
| Frequency of feeding-poor | 1.72 | 0.54 | 3.18 | .001 | 5.59 | 1.94 - 16.14 |

The reason why boys were more prone to weight faltering than girls does not appear to be related to feeding frequency according to the logistic regression analysis between sex and frequency of feeding. The odds of male sex predicting poor frequency of feeding was 0.91 (95% CI 0.5-1.66), but this was not significant at p=0.76. Illness occurrence in the boys and girls in the cohort was not significantly different, p=0.634.

### Comparing WHO weight-for-age z-scores, weight-for-length z-scores, headcircumference-for-age z-scores and length-for-age z-scores in the cohort

A Kruskal-Wallis test showed that there was a significant difference between the medians of the 4 categories of anthropometric z-scores, p=<.001. Table 4 showed that the median head circumference-for-age z-score was the lowest (−1.03), followed by the length (0.59). The highest was the median weight-for-age z-score (0.87).

**Table 4:** Medians of all the anthropometric z-scores.

| Groups | n | Median | Mean Rank |
| --- | --- | --- | --- |
| WTZ | 457 | 0.87 | 1095.29 |
| HCZ | 457 | -1.03 | 503.51 |
| LTZ | 457 | 0.59 | 958.92 |
| WLZ | 457 | 0.64 | 1100.29 |
| Total | 1828 | 0.23 |  |

Post hoc Test

A Dunn-Bonferroni test was used to compare the groups in pairs to find out which was significantly different. The Dunn-Bonferroni test revealed that the pairwise group comparisons of WTZ - HCZ, WTZ - LTZ, HCZ - LTZ, HCZ - WLZ and LTZ - WLZ had an adjusted p-value < 0.05 and thus, based on the available data, it can be assumed that these data and groups are significantly different in each pairs. However, there was no difference between the median weight-for-age z-score and median weight-for-length z-score, adjusted p-value was 1.

### Relationship between frequency of breastfeeding and the weight-for-age z-score

A linear regression analysis was performed to examine the influence of the variable frequency of breastfeeding in the previous 24hrs (FF24) on the variable weight-for-age z-score (WTZ).

Model Summary: The regression model showed that the variable FF24 explained 5.33% of the variance from the variable WTZ. An ANOVA was used to test whether this value was significantly different from zero. Using the present sample, it was found that the effect was significantly different from zero, F=25.62, p = <.001, R2 = 0.05.

Regression coefficients-The following regression model is obtained:

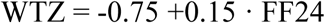

When FF24 was zero, the value of the variable WTZ was −0.75. On the reverse, when WTZ is 0, the value of the variable FF24 was 9.56.

If the value of the variable FF24 changes by one unit, the value of the variable WTZ changes by 0.15. The p-value for the coefficient of FF24 is <0.001, which is significant, Table 5.

**Table 5:**
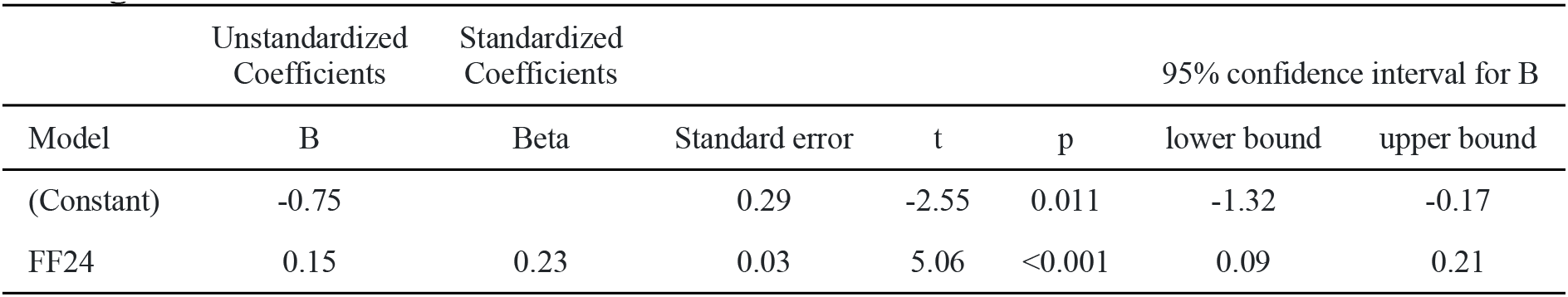
Linear regression analysis between frequency of breastfeeding predicting changes in weight-for-age z-score.

### Comparing the length of infants in the normal weight and weight faltering groups

The results of the descriptive statistics show that the normal weight infant group had higher values for the dependent variable Length (M = 60.5, sd = 4.29) than the weight faltering group (M = 53.15, sd = 2.65). The Levene test of equality of variance yielded a p-value of 0.011, which was significant and thus, there was no variance equality in the samples.

A two-tailed t-test for independent samples (equal variances not assumed) showed that the difference between normal weight and weight faltering groups with respect to the dependent variable length was statistically significant, t(19.4) = 10.92, p = <.001, 95% confidence interval [5.95, 8.77]. The effect size d was 2.7 (equal variances not assumed). With d = 2.7 there is a large effect.

### Comparing the head circumference of infants in the normal weight and weight faltering groups

The results of the descriptive statistics showed that the normal weight infant group had higher values for the dependent variable Head circumference (M = 38.32, sd = 1.29) than the weight faltering group (M = 37.05, sd = 1.4).

The Levene test of equality of variance yielded a p-value of 0.404, which was not significant and thus, there was variance equality in the samples. A two-tailed t-test for independent samples (equal variances assumed) showed that the difference between normal weight and weight faltering groups with respect to the dependent variable head circumference was statistically significant, t(455) = 3.97, p = <.001, 95% confidence interval [0.64,1.9]. The effect size d was 0.98 (equal variances assumed). With d = 0.98, there is a large effect.

### Type of illness experienced in the cohort

Fifty (10.9%) out of the 457 infants had been previously ill before presenting at the clinic, Table 6. Some had more than one type of diagnosis. 38 (76%) out of the 50 infants had respiratory problems, 10 (20%) had diarrhea illness, 2(4%) had malaria, 1 (2%) had sepsis and another 1(2%) had eye infection. 4 (8%) out of the 50 infants were hospitalized for their illnesses prior to presenting in the clinic.

The results of the descriptive statistics show that the group of infants who had experienced some form of illness prior to clinic visit had lower values for average frequency of breastfeeding (M = 8.62, sd = 2.28) than the group that had not experienced any ill-health (M = 9.97, sd = 1.75). The Levene test of equality of variance yielded a p-value of <.001, which was therefore significant and thus, there was no variance equality in the samples. A two-tailed t-test for independent samples (equal variances not assumed) was statistically significant, t(56.27) = −4.02, p = <.001, 95% confidence interval [−2.02, −0.68]. The effect size d was 0.6 (equal variances not assumed). With d = 0.6 there was a medium effect.

## Discussion

This study documented the prevalence of weight faltering (based on NICE static weight assessment method) among 457 infant cohort, that presented at the 10th week MCH clinic. Though the prevalence of weight faltering was slightly higher with the NICE criteria (3.7%) than that of the WHO criteria (2.8%), as expected, the level of agreement between the NICE weight assessment method and the WHO weight assessment method was significantly high at 93%. In addition,some risk factors of postnatal weight faltering were identified.

Of note, there was a further decline in the clinic attendance, from 529 at birth^14^, to 519 at 6 weeks,^15^ and then 467 at the 10th week clinic. 62 (11.7%) infants out of the original 529 participants recruited from birth, were absent at the 10th week MCH clinic. Another 10 babies were excluded from the statistical analysis due to incomplete data, and hence, only data from 457 babies were analysed. There is need to investigate and mitigate the factors contributing to decline in post-delivery clinic visits, as important medical conditions in both infant and mother may be missed in this critical phase of postnatal life.^14,15^

All the 457 babies were breastfeeding. Exclusive breastfeeding rate was still high at 96.3%, similar to the 96.4% obtained at 6weeks postnatal^15^for the same cohort. These exclusive breastfeeding rates were significantly higher than the 59.2% average recorded for infants 0-5months of age in the East African region, according to the 2022 Global Nutrition report.^16^

The significant factors in this study promoting infant weight faltering, despite high exclusive breastfeeding rate were male gender and low frequency of breastfeeding. As noted during the analysis, this observed sex-based weight faltering was not related to differences in the frequency of breastfeeding, nor to prior illness. In a systematic review, involving meta analysis of studies (2020) conducted in 30 countries globally, investigating boys’ increased tendency to undernutrition, reported boys to be more prone to wasting, underweight and stunting compared to girls in 59 out of the 74 studies investigated. The authors have suggested that this sex differences in undernutrition is more pronounced during the first 30months of life and then disappears thereafter, at older age groups. This was attributed to biologic (immune and hormones theories) and social factors.^17^

Despite the high prevalence of exclusive breastfeeding practices in this cohort, the frequency of daily breastfeeding was suboptimal in 10.3% of the infants. Infants that were poorly fed were 5.59 times more likely to falter in their weight compared to those that were adequately fed. For every unit increase in frequency of breastfeeding, the infant weight-for-age was likely to increase by 0.15 z-score, likewise, to maintain the infant’s weight-for-age at median of 0 z-score, caregivers may have to breastfeed on the average of 10 sessions per day.

The proportion of infants with weight faltering at 10 weeks post-delivery by static assessment was slightly higher than the 3.1% obtained at 6weeks in the same cohort.^15^ These rates were still significantly lower than the 50.4% reported amongst 4 months old Infants in an urban setting in Sri Lanka, where the exclusive breastfeeding rate at 4 months was 88%.^18,19^ Perhaps the lower exclusive breastfeeding rate in SriLanka, compared to that observed in this Rwanda study, could have contributed to the higher rate of infant underweight in Sri Lanka. Of the infant parameters measured in this study, the head circumference was the slowest in terms of growth, followed by the length, suggesting a need to include infant’s length and head circumference measurements in their routine clinic growth assessment and also to intensify exclusive breastfeeding agenda in this community. As of now, infant weight and occasionally the length is routinely monitored in the MCH facilities. Head circumference measurement is rarely performed in these well-child clinics.

It was also observed that adequate infant breastfeeding and weight thriving were accompanied with higher maternal depression scores, compared to those that were weight faltering and poorly fed, suggesting stress-associated experiences with breastfeeding and newborn care [maternal depressionogah].^20,21,22^ Rana et al. did mention that breastfeeding might be associated with some mental and physical ill health in these mothers,^3^ that might negatively feedback on exclusive breastfeeding and newborn care practices. There is a strong link between stressful life events (which may include difficulties with lactation and newborn care) and maternal depression. It appears these mothers were still struggling with these challenges at 10 weeks post-delivery since birth. This calls for a closer attention by the attending health workers, to the mothers, especially during the first 6 months postpartum, to enable them to achieve full exclusive breastfeeding and strenghtening of the lactational and mental support in these rural health facilities.

Even though these infants had recovered from their illnesses, as at the time of clinic visit, their previous 24hrs frequency of breastfeeding were significantly lower than those that had not been exposed to any ill-health prior to the 10^th^ week visit. This might suggest a lengthy period of recuperation from illness that was negatively impacting their feeding habits and hence growth parameters.

One of the limitations in this study was the loss to follow up of infants at the MCH clinic. Also, excluding small and sick babies from the study at birth, could have falsely depleted the infant weight faltering rate in the cohort. The authors recognise the variety of definitions of infant growth faltering employed in other studies, as there is no global consensus yet on the standard definitions of same for infants below the age of 6 months.

## Conclusion

This study emphasizes the importance of paying closer attention to both the practice of EBF and the daily frequency of breastfeeding and incorporating infant head circumference and length assessments in the routine growth monitoring programs in these rural MCH clinics. Lactational and mental support programs should be strengthened in the community, to assist mothers to achieve full exclusive breastfeeding easily. The factors contributing to decline in clinic visits need to be identified and mitigated. Home visits should be intensified for infants lost to follow up.

## Data Availability

All data produced in the present study are available upon reasonable request to the authors

## Author Contributions

The corresponding author (Dr Adenike Oluwakemi Ogah) conceived and designed the study, collected data and conducted data analysis, interpreted the results, and drafted the manuscript.

## Acknowledgements

The author is extremely grateful to the participants involved in this study, to the staff of Gitwe Hospital and clinic in Rwanda and to the research team.

